# Informing Pharmacokinetic Models with Physiological Data: Oral Population Modeling of L-Serine in Humans

**DOI:** 10.1101/2019.12.07.19011429

**Authors:** J. R. Bosley, Elias Björnson, Cheng Zhang, Hasan Turkez, Jens Nielsen, Mathias Uhlen, Jan Boren, Adil Mardinoglu

## Abstract

To determine how to set optimal oral L-serine (serine) dose levels for a clinical trial, existing literature was surveyed. Data sufficient to set the dose was inadequate, and so a (n=10) Phase I-A calibration trial was performed, administering serine with and without other oral agents. We analyzed the trial and the literature data using pharmacokinetic (PK) modeling and statistical analysis. The therapeutic goal is to modulate specific serine-related metabolic pathways in the liver using the lowest possible dose which gives the desired effect since the upper bound was expected to be limited by toxicity. In this paper, we first review relevant literature, describe the calibration trial and resulting data, and present the results of modeling from the trial. Serine is a non-essential amino acid that is nonetheless present at a base level in blood from both dietary sources and endogenous production. Serine is consumed by several pathways. A standard PK approach, in which a common model structure was selected using a fit to data, yielded a model with a single central compartment corresponding to plasma, clearance from that compartment, and an endogenous source of serine. The lack of intravenous data normally prevents independent determination of bioavailability and volume of distribution, however, under some assumptions about endogenous synthesis and use, values could be estimated. The model was poorly conditioned but did give consistent estimates. To improve conditioning, a parametric structure was changed to estimate ratios (bioavailability over volume, for example). Model fit quality was improved and the uncertainty in estimated parameters was reduced. Because of the particular interest in the fate of serine, the model was used to estimate whether serine is consumed in the gut, absorbed by the liver, or entered the blood in either a free state, or in a protein- or tissue-bound state that is not measured by our assay. The PK model structure was set up to represent relevant physiology, and this quantitative systems biology approach allowed a broader set of physiological data to be used to narrow parameter and prediction confidence intervals, and to better understand the biological meaning of the data. The model results allowed us to determine the optimal human dose for future trials, including a trial design component including IV and tracer studies. A key contribution is that we were able to use human physiological data from the literature to inform the PK model and to set reasonable bounds on parameters, and to improve model conditioning. Leveraging literature data produced a more predictive, useful model.

## Introduction

L-Serine (serine), a non-essential amino acid (AA), is *de novo* synthesized from glucose via 3-phosphoglycerate/3-phosphoserine and via interconversion of glycine. It can be obtained from diet, the degradation of dietary proteins and phospholipids and consumed in the production of pyruvate, amino acids (glycine, L-cysteine), and sphingosine and phosphatidyl serine. It also may be racemized to D-serine. Serine has attracted interest in the treatment of neurodegenerative diseases (de Koning et al., 2003), including Amyotrophic Lateral Sclerosis (Levine et al., 2017) and neuropathy (Gantner et al., 2019; Garofalo et al., 2011; Scherer, 2011).

As shown in Figure 1 (after (de Koning et al., 2003)), serine can be part of key metabolic pathways associated with proteins and carbohydrates, and supplemental serine is expected to affect hepatic metabolism. Mardinoglu *et al* have used Genome Scale Metabolic (GEM) modeling to infer that supplementation of serine may affect hepatic pathways that are relevant to the pathology of non-alcoholic fatty liver disease (NAFLD) and steatohepatitis (NASH) (Mardinoglu et al., 2014). Experimentally, a positive effect of serine therapy had been reported for alcoholic fatty liver in mice and rats (Sim et al., 2015). This led to a clinical trial of oral serine, showing a significant reduction in hepatic fat (assessed via imaging) with concurrent reduction of liver tissue markers (Mardinoglu et al., 2017). The trial data and further metabolic modelling analysis was used to generate a more comprehensive therapy approach, supplementation of metabolic cofactor formulation, of which serine is one of the component (Mardinoglu et al., 2017, 2019).

**Figure 1.**
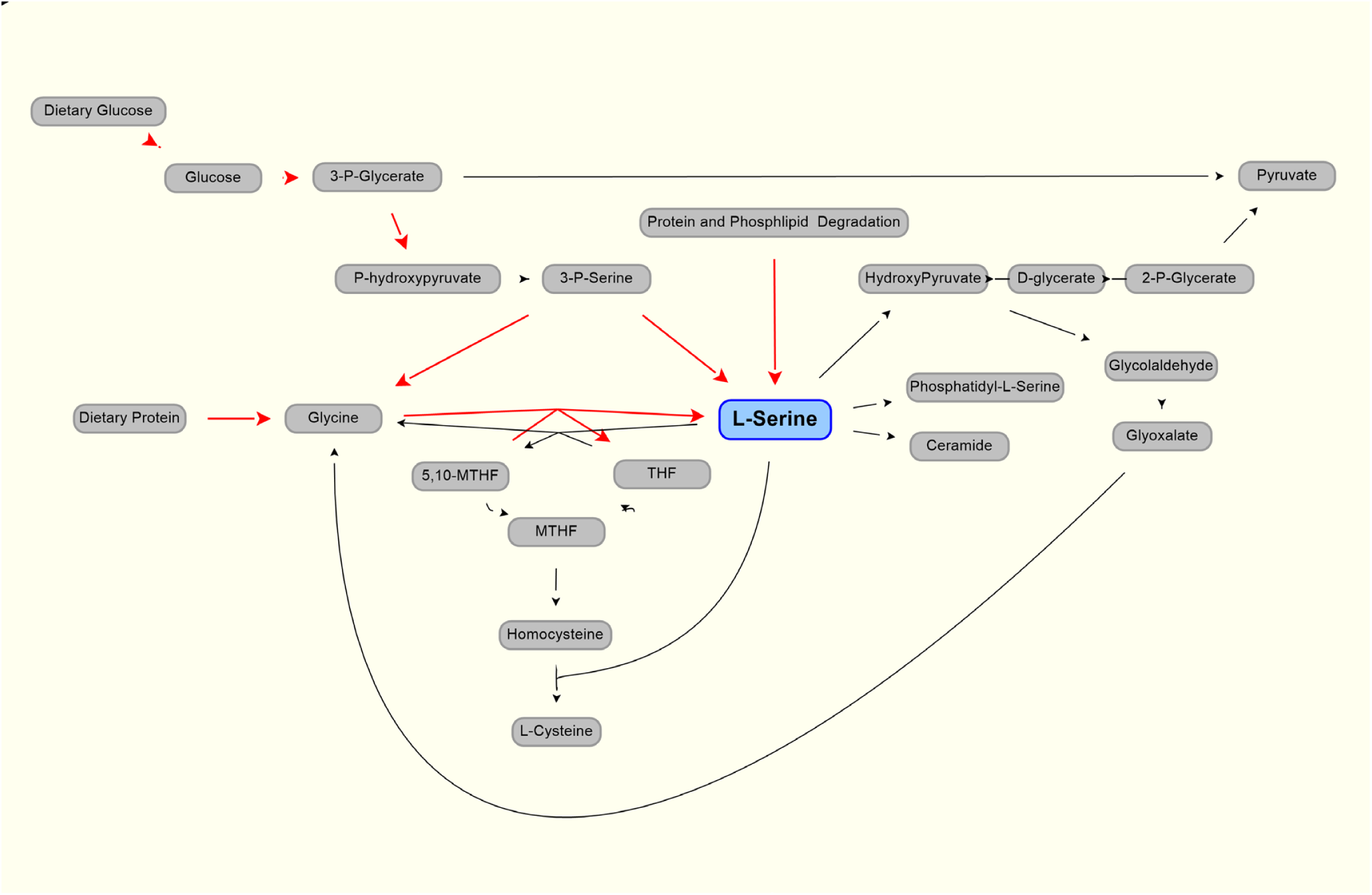
Production (red line) and use (black lines) of L-serine in the body. Modified from (de Koning et al., 2003)

Here, we aim to identify the optimal human dose of serine in this formulation to obtain the best therapeutic effect, and to avoid adverse effects in future clinical trials. Understanding clinical data and modeling the PK is complicated by the endogenous production of serine, predominantly (perhaps solely) from the kidney (Kalhan and Hanson, 2012; Pitts and MacLeod, 1972) leading to a measurable and significant baseline level in the blood.

### Review and Analysis of Literature Data

The human kidney extracts AAs (significantly, glutamine and glycine) and secretes other amino acids (in largest amounts, serine and alanine) (Pitts and MacLeod, 1972). Serine is extracted by the liver and other tissues including skeletal muscle (Felig, 1975). Exercise does not appear to affect serine concentration by more than five percent from the mean (Felig and Wahren, 1971). Glycine is the major precursor of serine, but glycine uptake is not adequate to account for all serine, and so glycolytic intermediates are also used by the body (Brosnan, 2003).

Published PK analysis of serine is sparse. (Pitts and MacLeod, 1972) reported values for volume of distribution of 199L and 360L for dogs weighing 20 and 28kg (10L/kg and 12.9L/kg, respectively). Baseline levels of serine in healthy subjects are published. A mean level (for five subjects) of 1.12 (with SEM of 0.046) mg/100ml, equivalent to 106 (4.3) umol/L was reported (Stein and Moore, 1954). These authors also cite a baseline serine level of 1.2 mg/100ml, equivalent to 114 umol/L, as being reported (Kurt Schreier and Hans Plückthun, 1950). A baseline serine value of 0.97 (0.07) mg/100ml, or 92.3 (6.3) umol/L was reported (Frame, 1958). Neis et al, (Neis et al., 2017) reported a value, measured in the radial artery, of 97.2 (4.7) umol/L and a value from the renal vein of 130 (8.4) umol/L, a positive difference (that is, kidneys releasing serine) of 32.8 umol/L. Using a standard value for kidney blood perfusion of 1.2 L/m (Calzia et al., 2005), and assuming that the assays capture all serine in the blood, we can estimate the net endogenous renal release of serine as approximately 4.2 mg/min or 252 mg/hr. A much higher estimate, which incorporates more detail, estimates serine production to be 150 umol//kg/h (Kalhan and Hanson, 2012), or about 1100 mg/hr. Endogenous production, at some level, clearly must be represented in the model. Detailed analysis of other amino acids show a time variation in concentration after meals in pigs (Stoll et al., 1998), and a similar variation in human serine production might be expected.

Wilcox et al, (Wilcox et al., 1985) show a mean level of serine of approximately 131 (SD of 18.5 SD) ug/ml for nonpsychotic control subjects. Interestingly, the baseline level shown for psychotics was 202.6 (SD 38) ug/ml, a difference that was reported as strongly significant (p<0.0001). These researchers determined an approximately 50% lower serine hydroxy methyl transferase (SHMT) enzyme activity in psychotic patients than in controls. The SHMT enzyme cleaves serine in the pathway synthesizing glycine from serine.

Wilcox et al, (Wilcox et al., 1985) also administered an oral bolus of 4mM/kg of serine to 12 actively psychotic patients, and 10 nonpsychotic subjects. This is equivalent to about a 29.5 g dose, for a 70kg subject. Using the oral dosing data, they found a range for serine elimination half-life of between 1.85 and 14.81 hours presumably due, at least in part, to SHMT level differences. There was a bimodal difference between active or previously active psychotic patients, and non-psychotic patients with no such history. The variability for psychotic patients was also much larger than for nonpsychotic subjects. The mean for non-psychotics was approximately 3.2 hours, with a standard error of 0.09 hours. The differences reported in SHMT may cause the longer half-life values and the higher baseline values. Clearly these factors must be considered in safe serine dosing in psychotic patients.

Serine is extracted from the blood by the liver in two major ways. There is a first pass clearance effect in which the liver extracts orally-dosed serine from the splanchnic circulation via the portal vein. In piglets fed with a mix of amino acids (Stoll et al., 1998) found that this first-pass metabolism in a fed state amounted to 58% of ingested serine. They also found that quantitatively more of the absorbed essential amino acids were catabolized than were incorporated into peptides and proteins. Hepatic extraction values for serine from arterial circulation of 24.1 and 55.0% were observed in rats receiving a 13% or 50% casein diet (Remesy et al., 1983). In roosters, a hepatic extraction value of 14% for serine was observed (Song et al., 2001)

In humans after 10-12 hours fasting, It has been reported that the small extraction of serine, that is the difference of 2.4 (5.1) umol/liter difference between portal vein and arterial concentration, was statistically insignificant (Felig and Wahren, 1971). In their study, the reported arterial serine concentration was 110.0 (11) umol/liter.

The mode of liver serine uptake is extraction of serine from the hepatic artery. Presumably this occurs in both fed and fasting states. In humans, it has been showed (graphically) a drop between arterial and hepatic vein concentrations of about 20 umol/liter, from a base of approximately 111 umol/liter in resting, postabsorptive (12-14 hour fast) state (Felig and Wahren, 1971). Again, using a standard value for hepatic perfusion of 1.4 liters/minute (Calzia et al., 2005), this would amount to 28umol/minute, or 3 mg/minute, or 180 mg/hr.

There is also extraction by muscle tissue. It has been reported that the arterio-femoral vein difference of serine for subjects in a postabsorptive state (10-14 hours fasting) to be 10.5 umol/liter with a standard error of about 4 umol/liter, with the base arterial level to be 110.0 +/- 11 umol/liter (Felig and Wahren, 1971). This work also gave an estimate of total blood flow to the legs, which was 0.9 (SEM 0.05) L/min. However, if a standard value for specific blood flow to muscle of 0.08 L/kg/min and the body is assumed to be 42% muscle, a 70 kg human will have muscle perfusion totaling 2.4 liters/minute (Calzia et al., 2005). If the serine extracted from all muscle averages the same 10.5 umol/liter, this gives an approximation of 25 umol/minute, or 2.6 mg/minute, or 156mg/hour.

The fasting values of 4mg/minute released by the kidney, and 3 and 2.6 mg/minute uptake into the liver and muscle, respectively, are approximations but the values can be used to check our model for gross error.

Garlick (Garlick, 2001) assessed hazards of increased oral amino acid intake and noted that data were sparse. Besides the paper by (Wilcox et al., 1985), which evaluated large doses (nearly 30g equivalent in a 70kg subject) in 10 control and 12 psychotic subjects and noted no adverse effects, a study of 4 actively psychotic schizophrenics by (Pepplinkhuizen et al., 1980) found that administering 2mmol/kg (which would be about 13 g total, for a 60kg subject) of serine to four female patients led to depersonalization in all, dysperception in 3, and euphoria and hallucination in one. Additional effects were noted. In all, the effects occurred about 5 hours after administration, and lasted 3-6 hours. None of the controls reacted to serine. This has implications when administering serine to trial subjects or patients with high baseline serine levels. It should be noted that serine does racemize to its enantiomer D-serine, and that D-serine is being investigated in schizophrenia – as a potential therapy (Kantrowitz et al., 2010, 2015), or as a therapy in combination with other therapies (Tsai et al., 1998) (Heresco-Levy et al., 2005).

(Levine et al., 2017) administered doses of 0.5, 2.5, 7.5, and 15g of serine twice a day in a trial with ALS (Amytrophic Lateral Schlerosis). While three of the 19 subjects died during the trial, in line with expected mortality for the cohort of ALS patients tested. Adverse effects which may be due to serine included one withdrawal due to bloating in one 15g b.i.d. patient.

## Results – Clinical Trial

Ten participants were recruited. All subjects were made aware of the risks inherent in the trial, and all agreed to participate. The study was performed in accordance with the Declaration of Helsinki and it was approved by the Ethics Committee at the Koc University, Istanbul, Turkey. Each subject gave written informed consent before participation in the study. The clinical trial was registered at ClinicalTrials.gov (with identifier: NCT03838822). Time limitations in recruiting limited trial subjects to males (future trials will include both sexes). One subject (subject 9) is a Type 1 diabetic and the other subjects were generally healthy. One subject (subject 10) had received the cocktail during the preceding two weeks, but that subject’s results were unremarkable and very close to responses of other subjects. Demographic data are summarized in Table 1. The protocol covered five days. All dosing was oral, in the morning after an overnight fast. On the first day, all subjects received 1 g of nicotinamide riboside. On day two, all subjects received 3 g of L-carnitine. On day three, all subjects received 5 g of N-acetyl cysteine. On day four, all subjects received a “cocktail” comprising the same dosing of NR, L-Carnitine, and NAC, and 20 g of serine. On day 5, a 20 g dose of serine alone was administered. Blood samples were taken every day (with number of samples per day being 2, 2, 1, 8, and 1 for days 1-5, respectively) and were analyzed for serine. Additional results are reported elsewhere. Here, we report only serine.

**Table 1.**
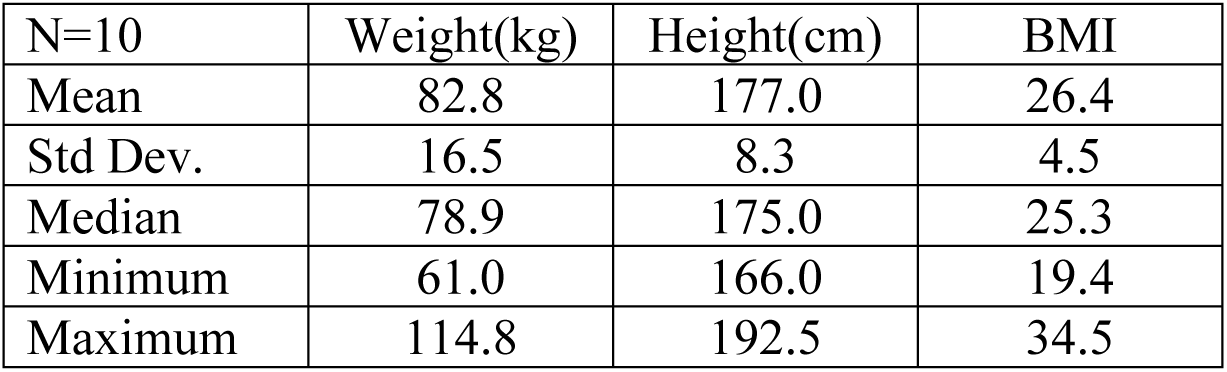
Trial subject demographic data

Serine was supplied as a powder. 20 g doses were prepared by weight and administered after mixing with 200 gr of water. There were no adverse effects reported by the participants during the trial. A trellis plot of the data for serine concentration over time is shown Figure 2. It is clear that a non-zero baseline value for serine is observed in all subjects. The average of the (pre-dose or baseline) serine values was 117 (6.7) umol/L, consistent with reported values given in the literature cited.

**Figure 2.**
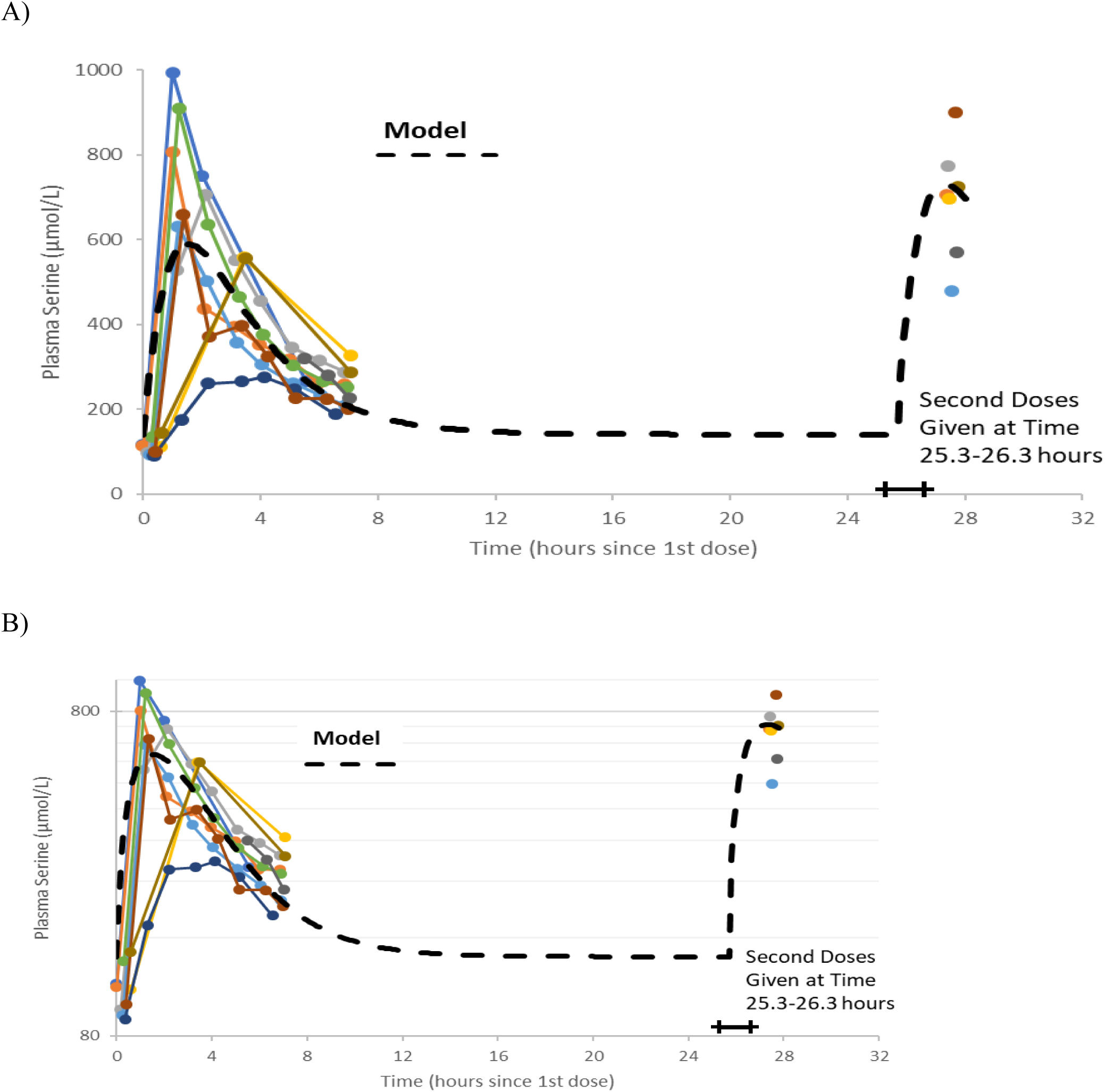
Plots of data, time-shifted so that the initial dose time is 0 hours. This resulted in the second dose time varying between 25.3 and 26.3 hours, shown as a horizontal bar in the figures. A naïve pooled model (dashed black line) is plotted with the individual data (colored dots and solid lines). A) Linear plot. B) Semi-logarithmic plot

### Modeling and Analysis

Figure 2A and 2B show the data on one plot, time-shifted so that the first dose was at time zero. The model trace shown in the figures is from a linear model, with mass action absorption of serine from a gut compartment to a plasma compartment subject to a bioavailability, mass action clearance from the plasma, and a constant endogenous biosynthesis being added to the plasma compartment. This is represented graphically in Figure 3. The modeling equations used were as follows. First, the amount of serine in the gut dose compartment, D, is

**Figure 3.**
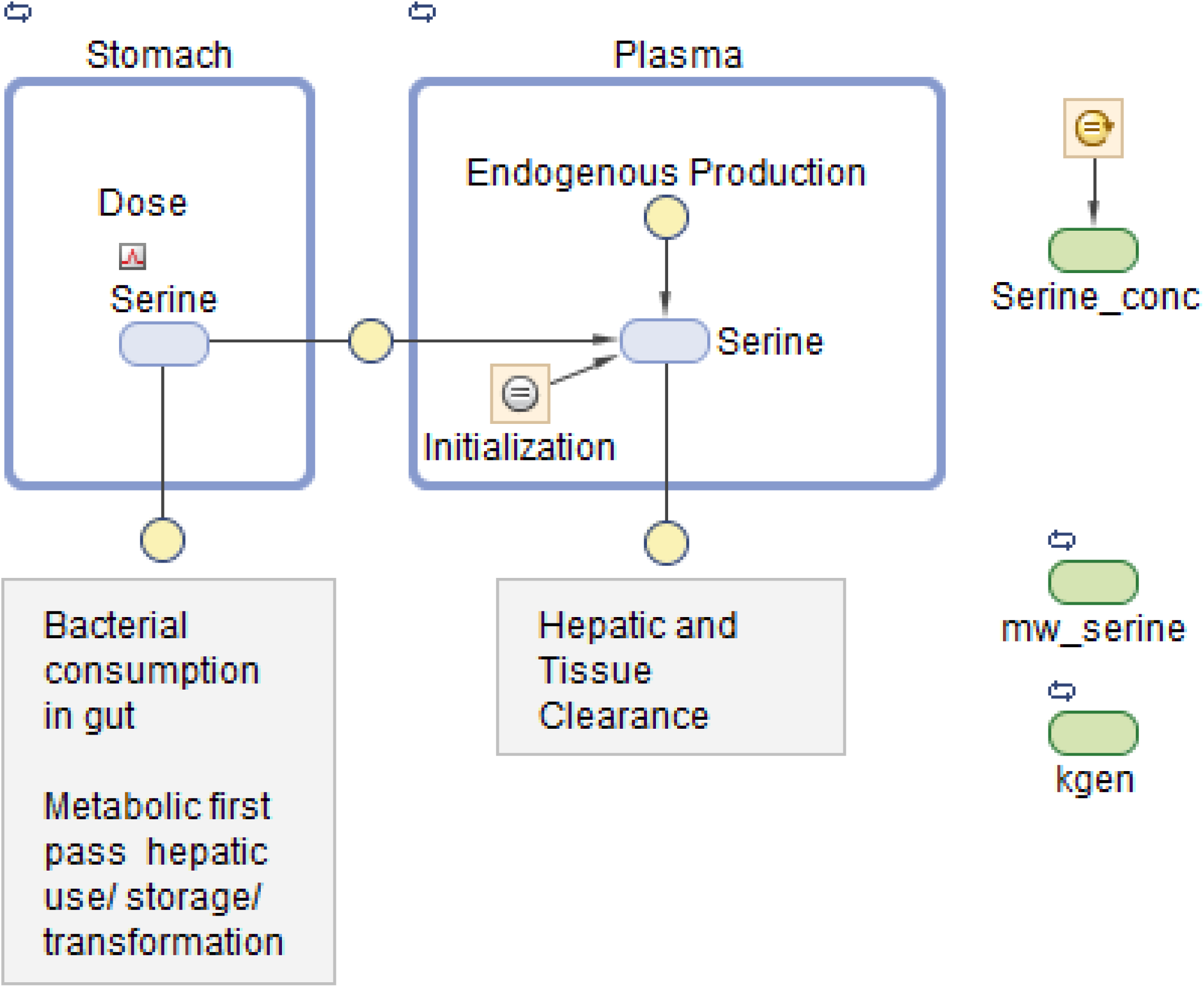
Graphical model representation from SimBiology(R)

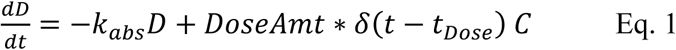

Where d is the Dirac delta function and DoseAmt is the bolus dose quantity.

The equation for Serine concentration, C, with five adjustable parameters, (V_D_, F, k_abs_, k_gen_ and CL) is

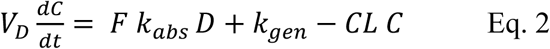

Where C denotes serine concentration, V_D_ is the apparent volume of distribution of serine, F is bioavailability, k_abs_ is a rate constant for absorption, k_gen_ denotes a constant endogenous serine production rate, CL is serine clearance.

One modeling assumption that should be highlighted is that endogenous serine production is assumed constant. One conclusion is that the administered dose of serine is virtually cleared within 12 hours.

Note that without a pre-existing value for the volume of distribution or bioavailability, this form of equation is usually poorly conditioned and any set of parameters (V_D_, F, k_abs_, k_gen_ and CL) that fits data could be scaled by an arbitrary amount to fit the data equally well. Nonetheless, a least-squares fitting process using naïve pooled technique did converge to a volume of about 1 liter. We know that this is likely not correct, as (Pitts and MacLeod, 1972) estimated volumes for dogs of approximately 160 liter. Parameter values are shown in Table 2. The individual data, and the naive pooled model results are plotted in figures 2A and 2B.

**Table 2.** Naive pooled fitted parameters for the poorly conditioned model shown in Figures 2A and 2B

| Dose | ka | F | CL | baseline | V |
| --- | --- | --- | --- | --- | --- |
| - | Absorption | bioavailability | Clearance | concentration | Volume |
| 20 | 0.829 | 0.00452 | 0.446 | 1.474 | 0.936 |
| g | 1/hour | dimensionless | L/hour | mg/100ml | liter |

The terminal clearance coefficient (CL/V_D,_ i.e. the rate of endogenous generation) is 0.48/hr. This is equivalent to a half-life of 1.45 hours, a bit lower than reported by (Wilcox et al., 1985). The rate of endogenous generation is calculated (CL * baseline) as 6.6 mg/hour, significantly lower than discussed above. This assumes a volume of distribution of about 1 liter, though, which is significantly at odds with the 199 and 260 liter values for dogs cited by (Pitts and MacLeod, 1972).

In addition to the non-physiological value for volume, the previous fit uses a naïve pooled-date approach and hence does not represent any subject very well, as the averaging process distorts absorption and clearance parameters and yields unrepresentative values for, for example, predicted maximum concentration, or Cmax. This motivated the use of population modeling.

The model may be modified to improve conditioning and to allow more detailed representation of the physiology discussed above so as to allow the use of a wider range of data. Also, one desired use for the model was to simulate clinical trials. Models that represent an average patients, but not any individual patient are not adequate for these tasks.

Model fitting without prior knowledge of volume (or bioavailability) will require modification to equation 2 to improve conditioning as shown:

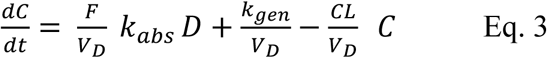

This reduces the degrees of freedom by one, as the fitting must solve only for four adjustable constant parameters, (F/V_D_, k_abs_, k_gen_ /V_D_ and CL/V_D_).

To generate parameter estimates and distributions, and to understand parametric distribution and confidence intervals for model prediction, we proceeded to use population pharmacometric approaches to modeling (Mould and Upton, 2013; Owen and Fiedler-Kelly, 2014; Upton and Mould, 2014, 2014).

The model was first implemented in SimBiology (MathWorks Inc., Natick MA, USA). SimBiology allows a graphical approach to model building. The picture of the model has underlying mathematics, and fitting and data manipulation can be done using integrated software. The model is pictured in Figure 3.

Before applying population methods, a first step in fitting this model was to fit each subject individually. This was done using the nonlinear least-squares (lsqnonlin) routine. The mixed (constant plus proportional) error model was found to give the best results. The results are shown in Figure 4 and parameter estimates are given in Table 3.

**Table 3.** Parameters from individual fit.

|  | kabs | F/V x 1000 | CL/V<sub>D</sub> | k<sub>gen</sub>/ V<sub>D</sub> |
| --- | --- | --- | --- | --- |
| Mean | 4.67 | 4.94 | 0.346 | 35.8 |
| Std Dev | 3.33 | 0.96 | 0.087 | 11.0 |
| Minimum | 0.51 | 3.47 | 0.198 | 20.6 |
| Maximum | 15.6 | 6.51 | 0.505 | 54.5 |
| Units | 1/hour | 1/liter | 1/hour | umol/(hr*liter) |

**Figure 4.**
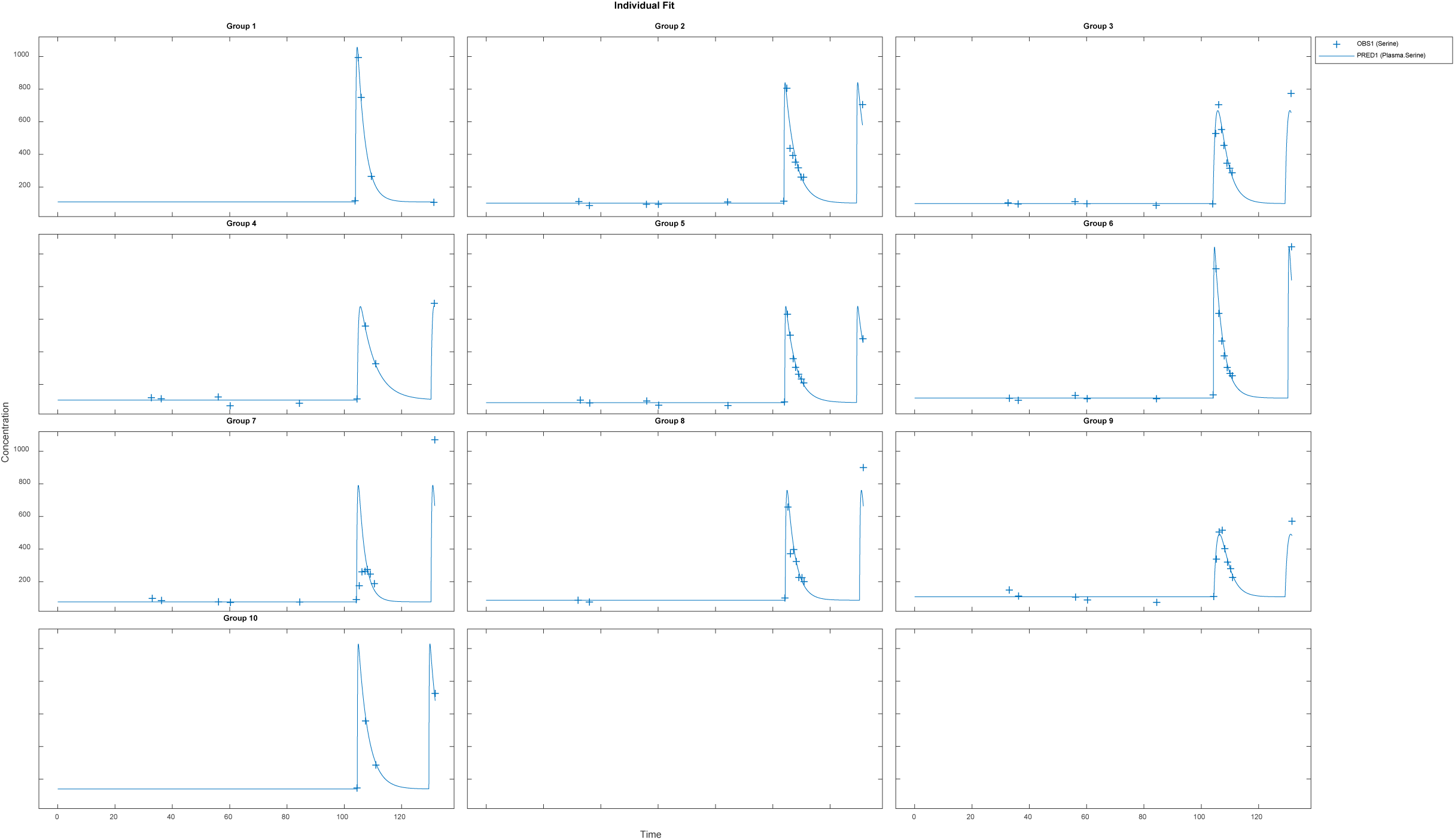
Results for fitting each subject individually, using a nonlinear least-squares routine with combined (constant plus proportional) error model. Clinical data (+) and model simulation (line)

Again, the mean values given cannot be guaranteed to actually represent any individual. That is, using these parameters in a simulation may result in an outcome that is not physiologically feasible or reasonable. A way to achieve mean parameters that are representative is to use a population approach. The nlmefit (non-linear mixed effect modeling) routine in SimBiology (MathWorks Inc, Natick, MA) was used using a combined (constant plus proportional) error model. The approach yielded the fits shown below:

The resulting parameters are given in Table 4. What is striking is just how small the variabilities for several parameters are. Most of the variability is in the bioavailability (F/V) parameter. The parameters were estimated as following a lognormal distribution, and so the parameter values estimated as the natural logarithm of the parameter, with standard errors are in logarithmic terms. To give ranges in meaningful units, the exponential of the nominal value and the nominal value plus and minus one standard error were calculated. Model results are plotted in Figure 5.

**Table 4.** Parameters from population fit using SimBiology® nlmefit with proportional error model

|  | kabs | F/V x 1000 | CL/V<sub>D</sub> | k<sub>gen</sub>/ V<sub>D</sub> |
| --- | --- | --- | --- | --- |
| Population | 2.88 | 4.29 | 0.288 | 29.6 |
| Pop - SEM | 1.88 | 3.96 | 0.266 | 27.0 |
| Pop + SEM | 4.42 | 4.64 | 0.312 | 32.3 |
| Minimum | 2.88 | 3.47 | 0.288 | 29.6 |
| Maximum | 2.89 | 4.87 | 0.288 | 29.6 |
| Units | 1/hour | 1/liter | 1/hour | umol/(hr*liter) |

**Figure 5.**
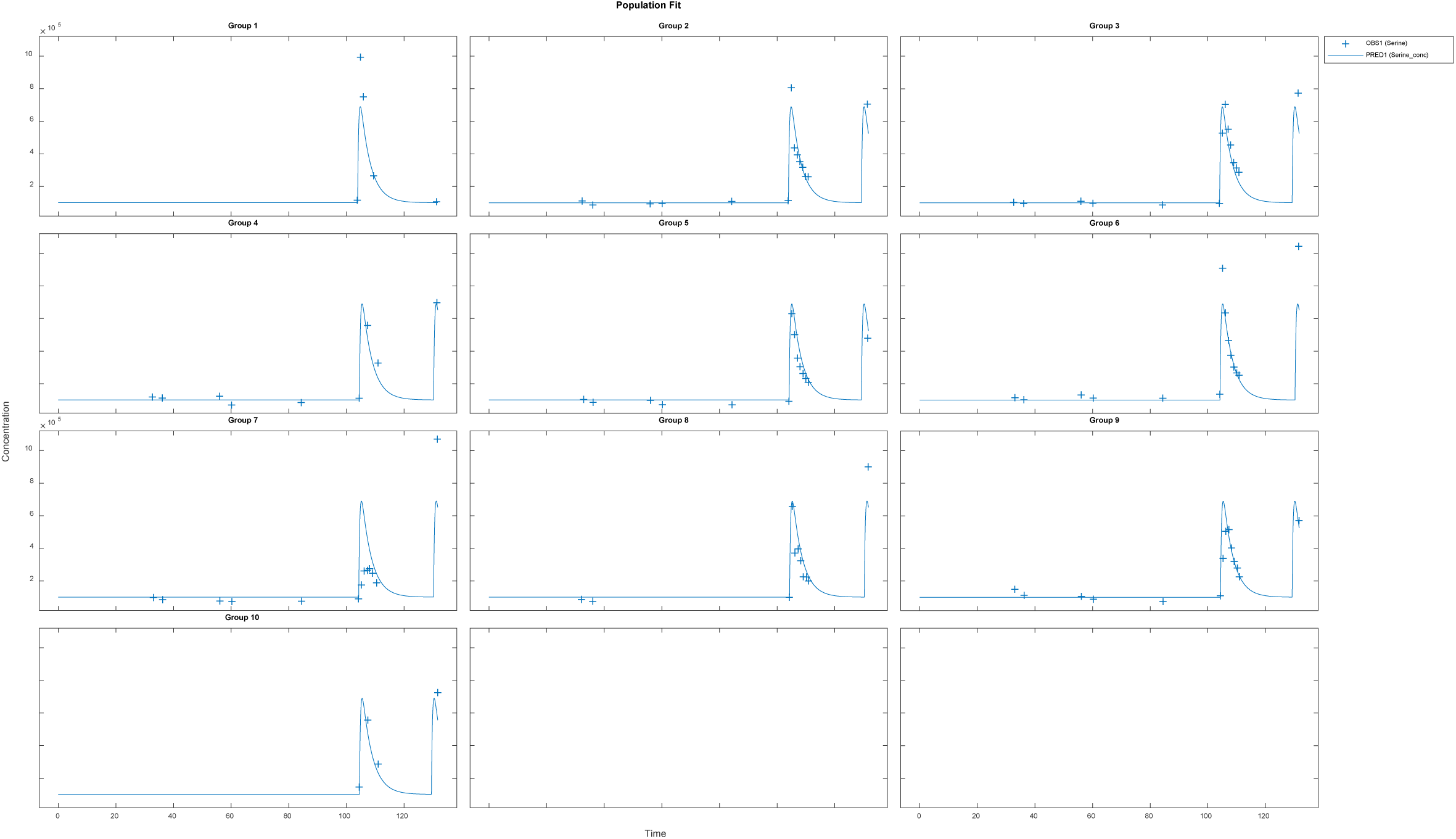
Results for fitting all subjects using a population fit and a combined (constant plus proportional) error model. Clinical data (+) and model simulation (line)

**Figure 6.**
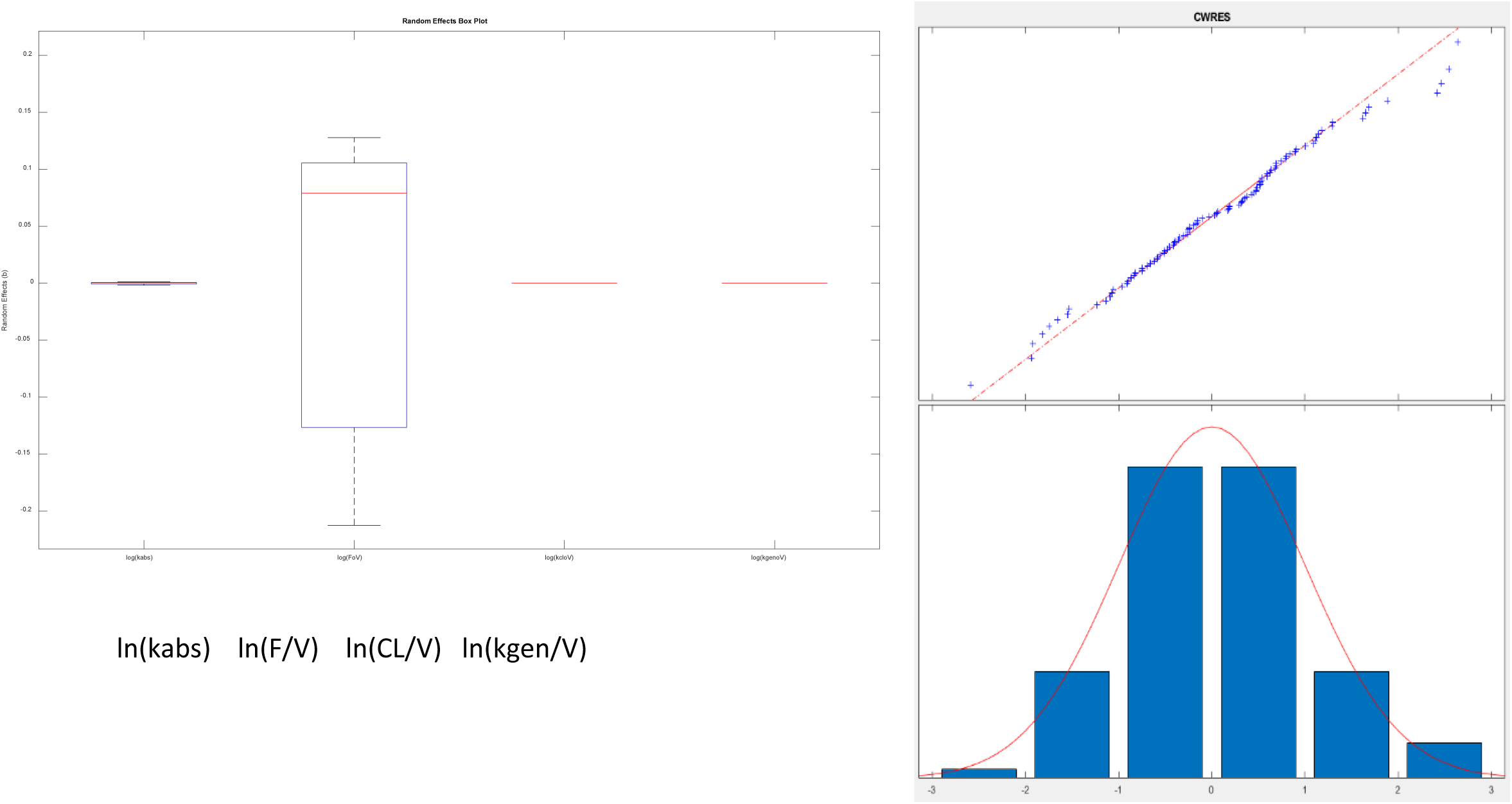
A) (above) Box and whisker plot of estimated parameters from population fit. Most variability is in the F/VD parameter. B) (at right) Plot of residuals to evaluate normality using a combined (constant plus proportional) error model. The combined model gave improved normality in the “tails” at both low and high values shown here, as opposed to a constant error model.

To check these results, the same model was programed in NONMEM® (ICON Plc, Ellicott City, MD). Parameter distribution was implemented as lognormal. The proportional error model gave nearly the same results as the mixed model, and so proportional error was used. The results for the proportional error model are given in Table 5.

**Table 5.** Population fit parameters from NONMEM.

|  | kabs | F/V x 1000 | CL/V<sub>D</sub> | k<sub>gen</sub>/ V<sub>D</sub> |
| --- | --- | --- | --- | --- |
| Population | 2.75 | 4.58 | 0.312 | 32.1 |
| Eta value | 0.921 | Small (FIXed) | 0.0135 | Small (FIXed) |
| Units | 1/hour | 1/liter | 1/hour | umol/(hr*liter) |

The Eta values that are shown as small were so small as to not allow noticeable variability (no change in 3 significant figures) and could be fixed in the estimation scheme. We note that the variability in the NONMEM parameters yields qualitatively different result than observed with nlmefit. One plausible explanation is that the two approaches found different local minima (Ricardo Paxson and Florian Augustin of MathWorks Inc, personal communication) as parameter variance estimates in nlmefit initially are set to zero, while in NONMEM®, these values may be initialized via the OMEGA vector or matrix.

We evaluated whether a second compartment improved the model. The improvement in the log-likelihood-based NONMEM metric (967.5 vs 979.785 for the one compartment model) lead to a difference of 12.2 which is more than the threshold of 5.99 (Dennis Fisher and Steven Shafer, 2007) and would justify the addition of two additional parameters (rate-law constants k23 and k32). However, the fitted value of these parameters were four orders of magnitude less than clearance and absorption rate constants and resulted in other parameter estimates changing only in the third decimal place with a change in predicted values that was trivial in relation to serine concentration. The additional compartment complexity was rejected on this basis.

We evaluated weight and height as potential covariates. Of the two parameters that showed variation between subjects, neither was shown to be correlated in any way to either metric. Likewise, there were no meaningful trends observed between other metrics (e.g. residual values) and weight and height. Our final model did not include covariates.

## Discussion

By using a model that was consistent with physiology, a broad range of published data for serine was able to be exploited. Model structure was improved, and we were able to build confidence in some fitted parameters (such as endogenous production rate). Animal extraction data (Neis et al., 2017) were used to estimate bioavailability (∼60%) and ultimate hepatic serine uptake fraction (75%). We were able to reject reported volume of distribution values from animal data (Pitts and MacLeod, 1972) as inappropriate for scaling to humans. Without intravenous data, a value for volume of distribution was estimated for serine (∼140L).

The dosing of 20 g QD in normal patients did not elicit any adverse effects in our clinical trial. The work of Wilcox et al. (Wilcox et al., 1985) noted no adverse effects in normal and psychotic subjects for a dose of 4 mM/kg, which is equivalent to 0.424 g/kg, or a 29.7 g dose for a 70 kg subject. Wilcox et al. (Wilcox et al., 1985) did note a higher baseline serine value, and lower SHMT activity, in psychotic subjects, and (Pepplinkhuizen et al., 1980) did observe schizophrenia-like psychoses induced in four active female schizophrenics (2 mM/kg, or 0.212 g/kg, equivalent to 12.7 g dose in a 60kg women). This suggests, but does not prove, that a dose of 20g or less may be tolerated in those who are not actively psychotic. Given that biopsy and/or tracer studies for SHMT activity are intrusive, a baseline serine value should be considered as an inclusion/exclusion test for future trials.

With that said, the 75% overall hepatic extraction with a 20 g dose lead to 15 g absorbed by the liver. Averaging this over 24 hours yields 10.4 mg/minute – a bit more than twice the current value of endogenous production, from clinical measurements. Thus, a 20g daily dose is expected to approximately triple hepatic serine uptake. Alternately, an approximately 12 g dose twice a day could be considered, due to the quick clearance of serine.

Systems modeling allowed the use of a broader set of data to expand our understanding of trial results. Reported parameters were used to rule out our first naïve pooled model, and to bound parameters in our population PK model. The population PK model was then used to inform our dosing decision for an upcoming trial. The modeling also highlighted the need to determine volume of distribution and bioavailability.

What was not considered was the variation of serine release and uptake during the day. We believe that this variability is inconsequential for our purpose of dose-setting, but the model could be augmented should this variation be relevant. We did not include the extraction of serine by the brain, but this could be easily analyzed using a modification of the current model and relevant trial data, for example using the results of (Levine et al., 2017), and a completed trial of serine in neuropath (Fridman et al., 2019).

A key interest in dose determination for serine is estimating the amount absorbed by the liver. Hepatic extraction will be both first-pass, from splanchnic circulation into the hepatic portal vein, and also system extraction from arterial circulation. Supposing that the 24.1 or 55 percent extraction range observed by (Remesy et al., 1983) in rats applies to humans, and assuming other use (for example, bacterial consumption in the gut) is negligible, this would imply a standard oral bioavailability values of 75.9 and 45%. Using the fitted value for F/V of 0.00429 gives values of volume of distribution of 177 and 104L. The question is whether these numbers and ranges are physiological.

It has been reported that the values for volume of distribution of 199L and 360L for dogs weighing 20 and 28kg (10L/kg and 12.9L/kg, respectively, must be scaled (Pitts and MacLeod, 1972). One source gives blood volume in dogs as 79 ml of blood per kg of body weight (Courtice, 1943). From this, we can calculate blood volume estimates of 1.580 and 2.212 liters. Allometric scaling principles suggest that these values might be scaled proportionally to body weight (Holford and Anderson, 2007), yielding (for both dogs) a value of about 5.5 liters of blood for a 70kg human, only about 10% higher than the commonly used value of 5 liters. The volume of distributions from dogs can be scaled proportionally to weight, yielding 700 and 900 liters. Using the average value of 800 liters, and 0.00429 for F/V gives an infeasible bioavailability of 340%. This suggests that the volume of distribution of serine reported for dogs (Pitts and MacLeod, 1972) is not appropriate or useful for humans, or at least that allometric scaling was not correct. Because of this, and because we suspect that extraction may be a more intrinsically conserved between species, the hepatic extraction values reported in rats (Remesy et al., 1983) were used to generate bioavailability. A bioavailability of about 0.6 (roughly the average between the 0.759 and 0.45 values in rats) yielded an effective volume of distribution of about 140 liters.

Of key interest is the fate of the approximately 60% of serine that is not initially absorbed by the liver. Recall the estimate above that the liver consumed about 3mg/minute, and the muscle tissue about 2.6 mg/minute. Assuming that these are the major consumers of arterial serine yields an estimate of arterial serine clearance of 46% by muscle and 54% by the liver. These values suggest that the liver extracts about 73% (40% initially, and another 46% of the 60% of serine that ends up in plasma) of ingested serine, with the rest going to muscle. This number is probably high, as some of the arterial serine is likely extracted by tissues other than muscle.

The population fit of endogenous serine generation was estimated to be 29.6umol/L/hr, as kgen/V. When multiplied by the estimated volume of 140L, this gives 440.0 mg/hour. This must be compared with the 252 mg/hour estimated from (Neis et al., 2017), and the 1110 mg/hour estimated from the work of (Kalhan and Hanson, 2012) which was felt to be high. Given differences in protocols, subjects, and variability of measurements, these value does not compare unfavorably, and provide some confidence in the model analysis.

If 440mg is generated endogenously every hour, and the liver accounts for the uptake of 54% of that amount, normal liver uptake of serine is about 6 g of serine per day. An oral dose of 8 g/day (6 g/73% liver utilization for oral dosing) could approximately double liver uptake, and a dose of 16 g/day could be expected to triple liver uptake, provided the extraction values considered here did not change. Earlier experiments with once-per-day 20g dosing (Mardinoglu et al., 2017) gave good results in some subjects, weaker responses in others. Noting that daily dosing up to about 30 g have been administered to normal subjects without adverse effects, and desiring a stronger effect suggested that a prudent upper bound would be a total of 25g per day. Given that the serine is apparently cleared quickly and a sustained metabolic effect is desired, the recommended 25 g dose could be administered as 12.5 g, twice a day. This was the recommendation for the upcoming trial. A more comprehensive clinical trial is planned and will be executed. A portion of that trial will include intravenous dosing to allow a better determination of bioavailability. In addition, we expect to use tracer studies to confirm hepatic extraction values derived from the model.

## Data Availability

All data are available in the manuscript

## Acknowledgements

The authors are grateful to Mr. Ricardo Paxson and Dr. Florian Augustin of Mathworks, Inc. for their helpful criticisms and insight into the Simbiology nlme fitting routine.

